# Hemodynamic Oscillations in Mild TBI During Postural Change: An fNIRS Pilot Study

**DOI:** 10.1101/2025.09.25.25336687

**Authors:** Carsi Kim, Andrea Gomez Carillo, Ulas Sunar

## Abstract

We demonstrate that low-frequency oscillations (LFOs) in cerebral hemodynamics, measured by frequency-domain functional near-infrared spectroscopy (FD-fNIRS), reflect altered cerebral hemodynamic oscillations in mild traumatic brain injury (mTBI). In a pilot study of two mTBI and 13 healthy subjects undergoing head-of-bed positional changes, we analyzed total hemoglobin concentration (THC), oxyhemoglobin (HbO), and deoxyhemoglobin (Hb) dynamics using spectral and time-frequency analyses. mTBI measurements exhibited significantly larger postural changes in THC (ΔTHC = 9.49 µM) compared to controls (ΔTHC = 1.03 µM). LFO power was consistently elevated in mTBI across all slow bands (0.01-0.2 Hz), particularly in the Slow-5 band (0.01-0.027 Hz), suggesting dysregulated cerebral vasomotion. Continuous wavelet transform (CWT) confirmed persistent LFO amplification during and after postural transitions. These findings indicate that THC-based LFO measures may serve as early, non-invasive biomarkers of cerebral autoregulatory impairment in mTBI.

## 1. Introduction

Mild traumatic brain injury (mTBI) remains a diagnostic challenge due to the absence of objective biomarkers and the subjective nature of clinical symptoms such as headache, dizziness, and cognitive dysfunction. Unlike moderate or severe traumatic brain injury (TBI), mTBI often presents with a normal Glasgow Coma Scale (GCS) score, making diagnosis complex [1,2]. The complication in diagnosis is even more apparent in mTBI’s current diagnosis, stated as, any plausible mechanism of injury combined with clinical signs or symptoms, even in the absence of neuroimaging abnormalities [3]. This means that patients with a Glasgow Coma Scale (GCS) score of 15 out of 15 may still meet the diagnostic criteria if they experience acute symptoms or cognitive deficits but with no quantitative diagnostic metrics [4]. The lack of a universally accepted diagnostic standard leads to variability in clinical assessment, making subjective reports a significant factor in identifying mTBI cases [3,5–7]. Another large concern is cerebral ischemia, which when monitored with cerebral blood flow (CBF) and oxygenation-parameters reduces mortality rate from 44 to 25% [14]. Current modalities such as computed tomography (CT), magnetic resonance imaging (MRI), intracranial pressure (ICP), and transcranial doppler ultrasound (TCD) can diagnose TBIs but they are not optimal for continuous, or repetitive monitoring of mTBI.

A growing concern in mTBI is the potential for cerebral autoregulatory dysfunction, which may contribute to secondary injury and delayed recovery. While tools such as intracranial pressure (ICP) monitoring, transcranial Doppler (TCD), or advanced MRI can assess cerebral physiology, they are invasive, costly, or lack feasibility for continuous, bedside evaluation, especially in mild cases [8–10]. Functional near-infrared spectroscopy (fNIRS) is a non-invasive, portable modality capable of measuring cerebral hemodynamics in real time. It has been employed in neurological conditions to assess oxygenation, neurovascular coupling, and autoregulation [11–22]. Of particular interest are low-frequency oscillations (LFOs) in the hemodynamic signal, which occur below 0.2 Hz and are thought to reflect vasomotor activity and cerebral autoregulatory function [23–26]. Prior studies suggest that abnormalities in LFOs may serve as early indicators of vascular impairment in stroke, small vessel disease, and severe TBI [27–29].

fNIRS has proven effective in assessing cerebral oxygenation and autoregulation during resting state and brain injury conditions, evaluating consciousness levels, and monitoring cognitive function [11–22]. Recent advancements have demonstrated potential to track recovery patterns in TBI cases, identify focal vascular injuries, and widespread microvascular damage [1–3,30–32]. By applying frequency analysis to fNIRS hemodynamic data shows we can obtain low frequency oscillations (LFOs). LFOs are associated with vasomotion, which refers to the rhythmic oscillations in blood vessel diameter due to spontaneous vascular smooth muscle activity. LFOs have shown promise in assessing injury severity, recovery trajectories, and long-term outcomes across neurological and neurovascular conditions such as stroke, acute traumatic brain injury, small vessel disease, and sickle cell disease [13,16,28–30,33–46].

Particularly, LFOs in the 0.07-0.1 Hz range have been linked to cerebral autoregulation [23–26]. For example, fNIRS studies in ASD have also demonstrated altered low-frequency hemodynamic oscillation patterns, particularly in the Slow-5 (0.01-0.027 Hz) and Slow-4 (0.027-0.073 Hz) bands, highlighting the sensitivity of LFO-based analysis to neurovascular dysregulation [40]. This approach may offer a practical, non-invasive, and quantitative method for evaluating cerebral autoregulation in mild brain injury. Here, we assess whether fNIRS-derived LFOs, particularly in THC, differ between mTBI and healthy controls during postural change, can reveal altered cerebral autoregulatory responses in individuals with mTBI. By analyzing LFO spectral power during postural transitions (head-of-bed tilt), we aim to identify frequency-domain signatures that distinguish mTBI from healthy individuals.

## 2. Methods

### 2.1 Participants

The total number of subjects utilized for this project was 13 healthy, subjects and two mTBI subjects. The subjects consisted of 15 volunteers of which had a mean age of 24 years, ranging from 20-40 years old. There were eight males and nine females of which one female and one male met the criteria of a mTBI from a Glasgow coma score of 15 and 14, respectively. Both populations, mTBI and healthy, underwent the same ten-minute measurement protocol after a five-minute baseline stabilization period where no measurements took place. The Instutional Review Board of Wright State University approved this protocol, and participants consented to the measurements.

### 2.2 Data Analysis

A commercial Frequency Domain-functional Near-Infrared Spectroscopy (FD-fNIRS) system (OxiplexTS, ISS Inc) was used to acquire AC (Alternating Current) signals at multiple source-detector separations. The ASCII file output from the OxiplexTS system was imported into MATLAB R2024b for analysis. Only the AC intensity values for two wavelengths, 830 nm and 690 nm, and one source-detector (SD) distance at 3 cm, sampled at 50 Hz, was used for this analysis. AC data is less sensitive to room light, making it a preferred alternative to DC (Direct Current) intensity in noisy brightly lit environments [47]. The AC intensity values were converted into relative hemoglobin concentration values using the optical density and modified beer lambert law equations [48]. After oxyhemoglobin(HbO) and deoxyhemoglobin(Hb) concentrations were calculated, a bandpass filter with a range of 0.009–5 Hz was applied during frequency analysis to isolate physiological signals while eliminating high-frequency noise. A minute of data during the transition period was omitted to assess only the baselines. Using MATLAB’s Fast Fourier transform (FFT) was applied directly to the time-series data from the baseline conditions of elevated and the supine position to calculate power spectral density (PSD). This approach maximized frequency resolution, preserving spectral peaks of interest without introducing smoothing effects. As an additional evaluation for a more reliable and robust PSD measurement, MATLAB’s pwelch function was used. Welch’s method was applied to estimate the total power of LFOs during the steady-state conditions to show more accurate overall LFO bar plots. The FFT length (NFFT) was defined as the sampling rate divided by the desired frequency resolution. The window length was set equal to NFFT, with a 50% overlap between successive segments. The area under the PSD curves were grouped into LFO slow bands, Slow 5: 0.01-0.027 Hz, Slow 4: 0.027-0.073 Hz, Slow 3: 0.073-0.198 Hz. These sections of slow bands are hypothesized to relate to specific physiological parameters particularly seen from fMRI studies [49,50]. To analyze the time series with both time and frequency dynamics, a MATLAB’s continuous wavelet transform (CWT) was performed using the Morse wavelet. This technique allowed for time-frequency analysis, providing a view of transient LFO features during the entirety of the protocol [51–53]. An overview of the data analysis pipeline is summarized in Figure 1.

**Fig. 1.**
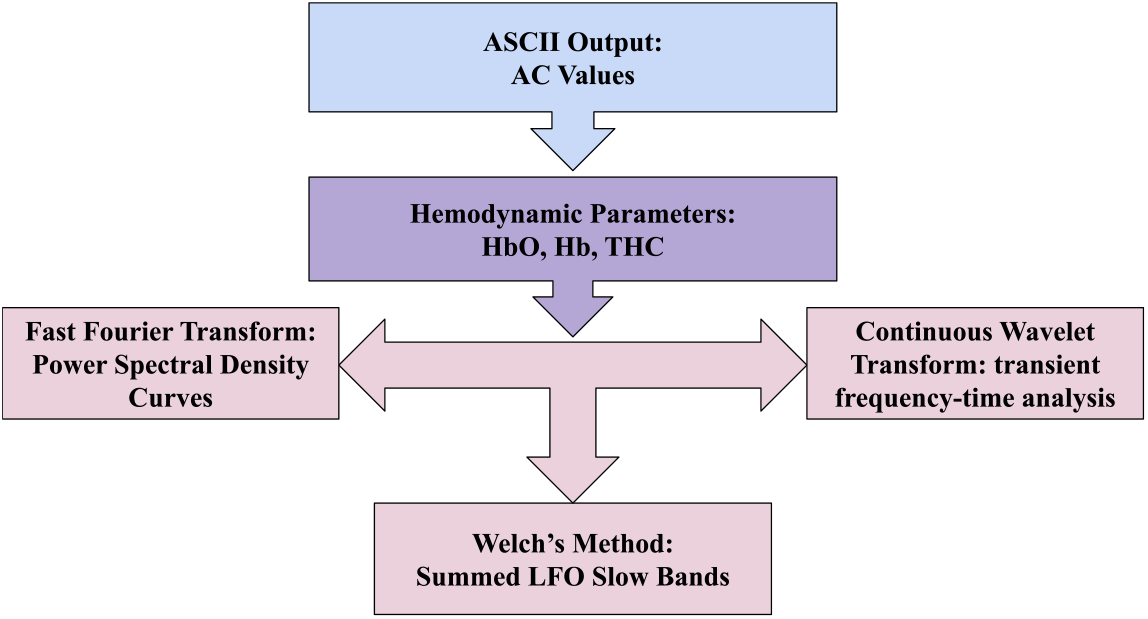
An overview of the data analysis pipeline. Starting at the ASCII output and AC (Alternating Current) value analysis to the calculation of hemodynamic parameters oxyhemoglobin (HbO), deoxy hemoglobin (Hb), and total hemoglobin concentration (THC). Then frequency domain analyses of fast Fourier transform and Welch’s method, and frequency-time analysis of continuous wavelet transform was done on the concentration data.

### 2.3 Experimental Protocol

Each participant underwent a standardized 15-minute head-of-bed (HOB) protocol. Before measurements, subjects were informed about the study procedures and provided written consent. The optical sensor was positioned on the left dorsolateral prefrontal cortex using a guided setup process. To ensure accurate placement, an EEG cap marked according to the international 10-20 system was utilized. The cap was fitted by measuring the nasion-to-inion distance with tape and positioning the Cz marker at the midpoint of this measurement. The cap was adjusted symmetrically on the subject’s head, and a small reference mark was placed above the left Fp1 position. After removing the cap, the sensor’s source optode was positioned slightly below the Fp1 marking to prevent interference with the signal. The entire sensor was then secured with an elastic band around the head. The placement setup is illustrated in Figure 2a.

**Fig. 2.**
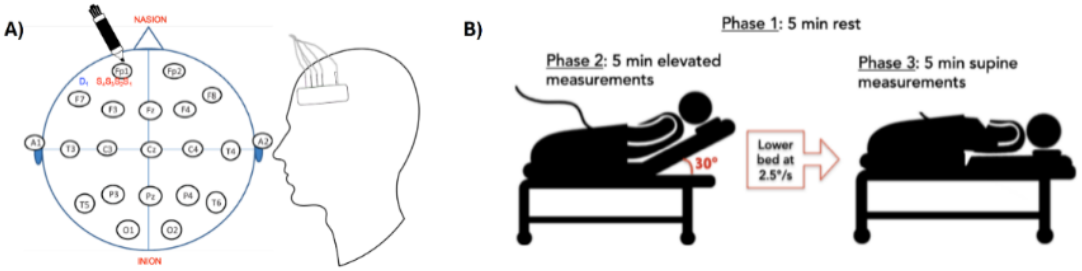
(a) Location of marking on the Fp1 position from the international 10-20 system and the placement of detectors and sources. Location of sensor on subject’s left dorsolateral prefrontal cortex. (b) The three phases of the head-of-bed (HOB) protocol: Phase 1 was baseline as the subjects rested in an elevated position, phase 2 measurements began at the elevated position for 5 minutes, phase 3 measurements continued at the resting supine position for five minutes.

The elevation changes during the protocol were facilitated using a motorized hospital bed to adjust the angle of elevation. Subjects were at an initial elevation of 30 degrees and asked to relax for five minutes to establish baseline conditions. After this resting period, a five-minute head elevated phase was recorded. Afterwards, the bed was lowered to a zero-degree angle at a controlled rate of 2.5 degrees per second, and recording continued for an additional five minutes. The three phases of the protocol are summarized in Figure 2b. This systematic approach ensured consistent and reliable measurements across all participants.

## 3. Results

An example of a subject’s time-series related to bed tilt going from elevated to supine is shown in Figure 3A. The general trend showed an increase in HbO and THC, with a decrease in Hb. The change between the averages of the two baseline concentrations was taken as Δ concentrations seen in Figure 3B. For the two baselines’ averages in concentrations were taken as seen in Figures 3C and 3D for supine and elevated respectively. For HbO, Hb, and THC, mTBI had a higher overall average concentration, in addition to THC showing the largest difference between the two groups in all three condition analyses.

**Fig. 3.**
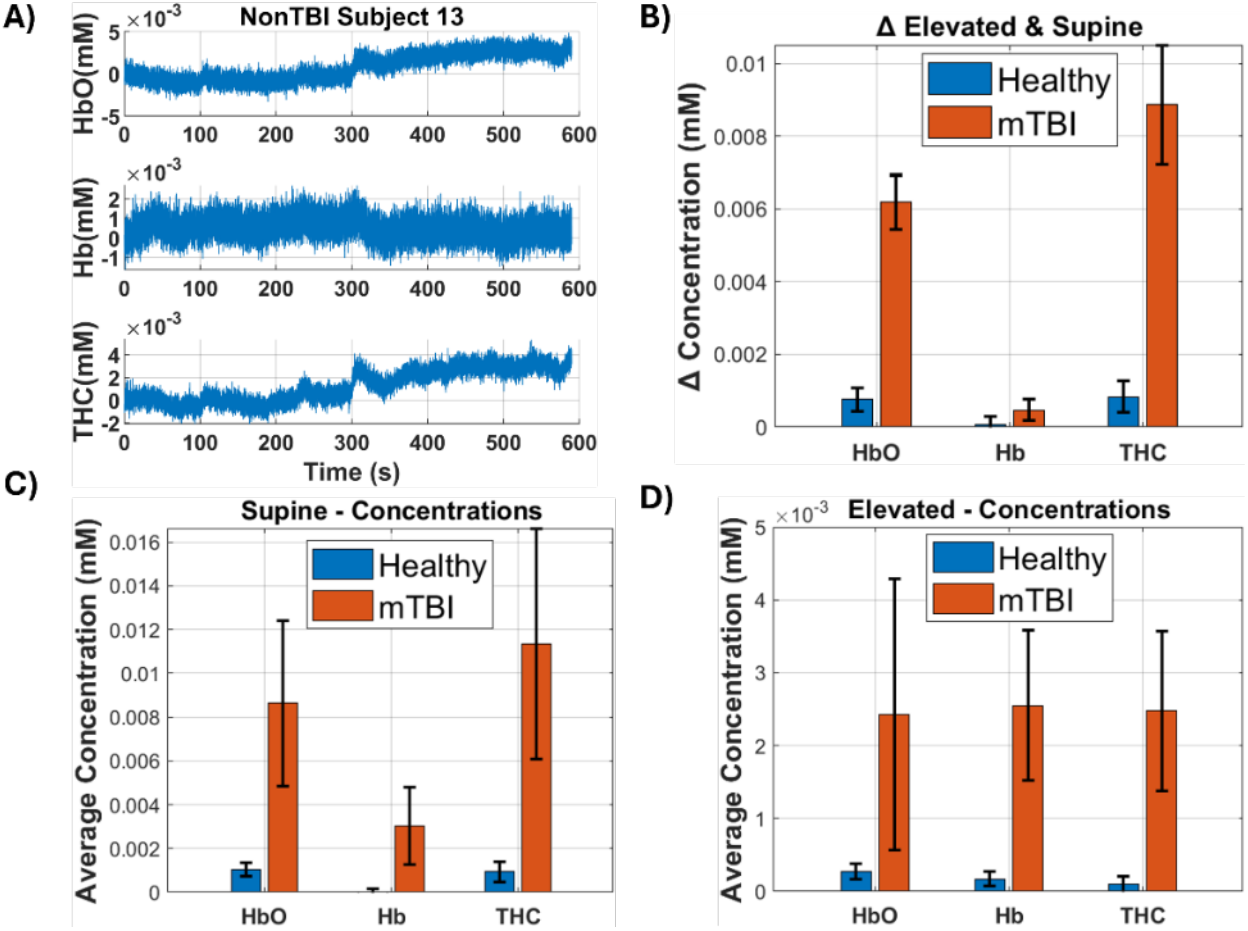
Averages for mild-traumatic brain injured (mTBI) and healthy relative hemoglobin concentrations, Deoxy-Hemoglobin (Hb), Oxy-Hemoglobin (HbO), and Total Hemoglobin Concentration (THC), of each baseline period were taken, omitting a minute of data in the middle of the data during the transition period during the bed tilting. (A) An example time series of hemodynamic parameters (HbO, Hb, and THC). (B) The change, Δ, in concentrations for mTBI and healthy for the hemodynamic parameters. (C) The average of mTBI and healthy supine HbO, Hb, and THC concentrations plotted into a box chart. (D) The average of mTBI and healthy elevated HbO, Hb, and THC concentrations plotted into a box chart.

In Figures 4, 5, and 6 the results of time-series and PSD analysis are shown for THC, Oxy-, and Deoxy-hemoglobin concentrations, respectively. In all three hemodynamic parameters, mTBI showed higher PSD in the lower frequencies compared to healthy. Using Welch’s method, PSD was also calculated to accurately portray the grouped LFO bands. PSD for each slow band range was summed within these bands Slow 5: 0.01-0.027 Hz, Slow 4: 0.027-0.073 Hz, Slow 3: 0.073-0.198 Hz. In mTBI, all LFO bands showed higher power compared to healthy. The average CWT was taken for each group to show the frequency and power over time. During the transition period (~300s) and more than a minute afterwards there is higher power in mTBI throughout the 0.15 - 0.009 Hz frequency range for all three hemodynamic parameters. This effect was most apparent in THC compared to HbO and Hb. Compared to Hb, HbO and THC show more similar trends in quantitative differences between the mTBI and healthy groups. Due to the small mTBI subject population number the data assumptions are little to no statistical significance, and results can be seen as preliminary or as a pilot study.

**Fig. 4.**
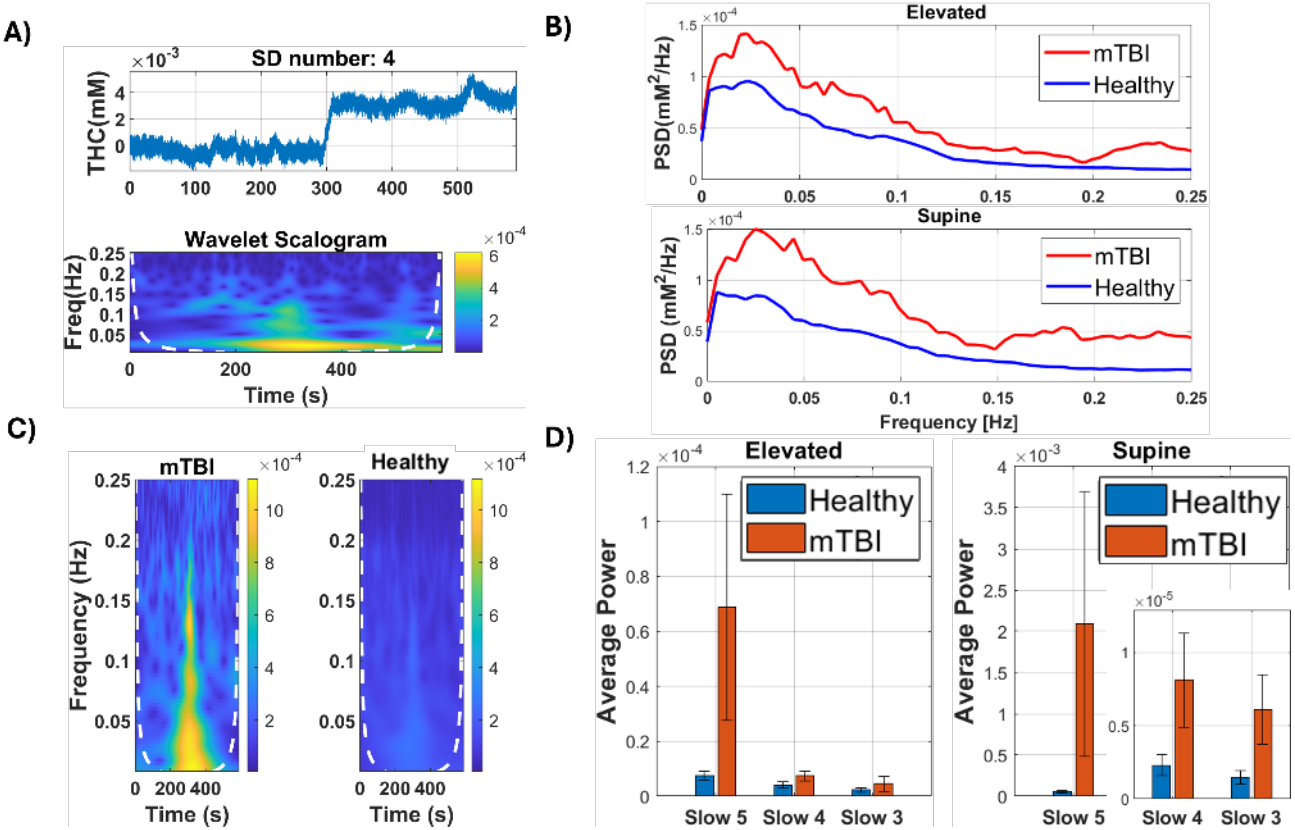
Analysis of Total Hemoglobin Concentrations (THC) in all subjects. (A) A single subject’s total hemoglobin concentration (THC) time series and the scalogram calculated from the continuous wavelet transform (CWT) are put side by side. This was done for every subject. (B) Healthy and mTBI elevated and supine positions are plotted in the frequency domain using fast Fourier transform. (C) The CWT plot shows a dashed white line showing the cone of influence (COI) that depicts areas where data interpretation is not accurate due to the stretching nature of wavelets. (D) Healthy and mTBI elevated and supine positions are plotted into summed bar plots processed using Welch’s method. Frequencies are grouped into slow bands, Slow 5: 0.01-0.027 Hz, Slow 4: 0.027-0.073 Hz, Slow 3: 0.073-0.198 Hz.

**Fig. 5.**
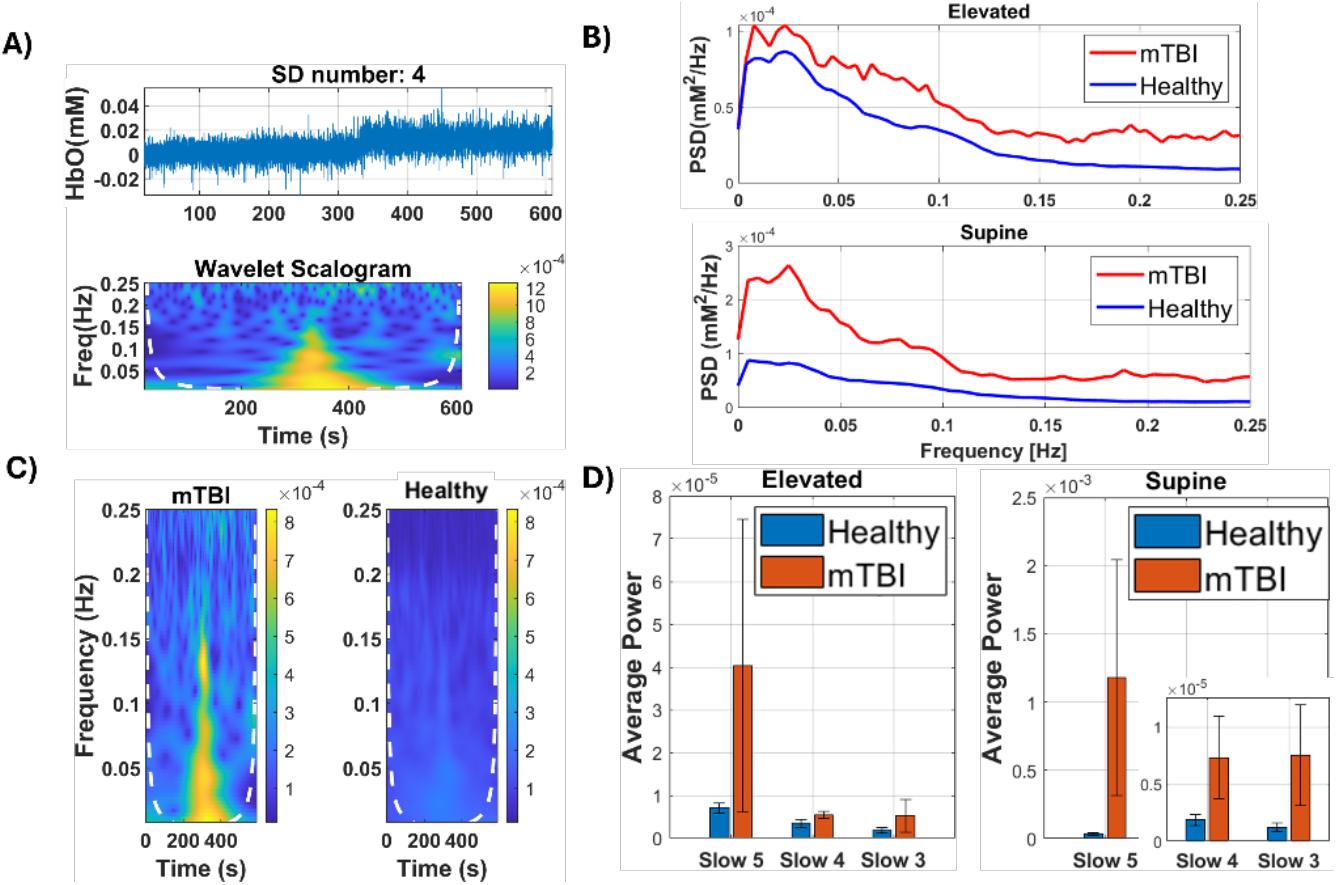
Analysis of Oxy-Hemoglobin (HbO) concentrations in all subjects. (A) A single subject’s HbO time series and the scalogram calculated from the continuous wavelet transform (CWT) are put side by side. This was done for every subject. (B) Healthy and mTBI elevated and supine positions are plotted in the frequency domain using fast Fourier transform. (C) The CWT plot shows a dashed white line showing the cone of influence (COI) that depicts areas where data interpretation is not accurate due to the stretching nature of wavelets. (D) Healthy and mTBI elevated and supine positions are plotted into summed bar plots processed using welch. Frequencies are grouped into slow bands, Slow 5: 0.01–0.027 Hz, Slow 4: 0.027–0.073 Hz, Slow 3: 0.073–0.198 Hz.

**Fig. 6.**
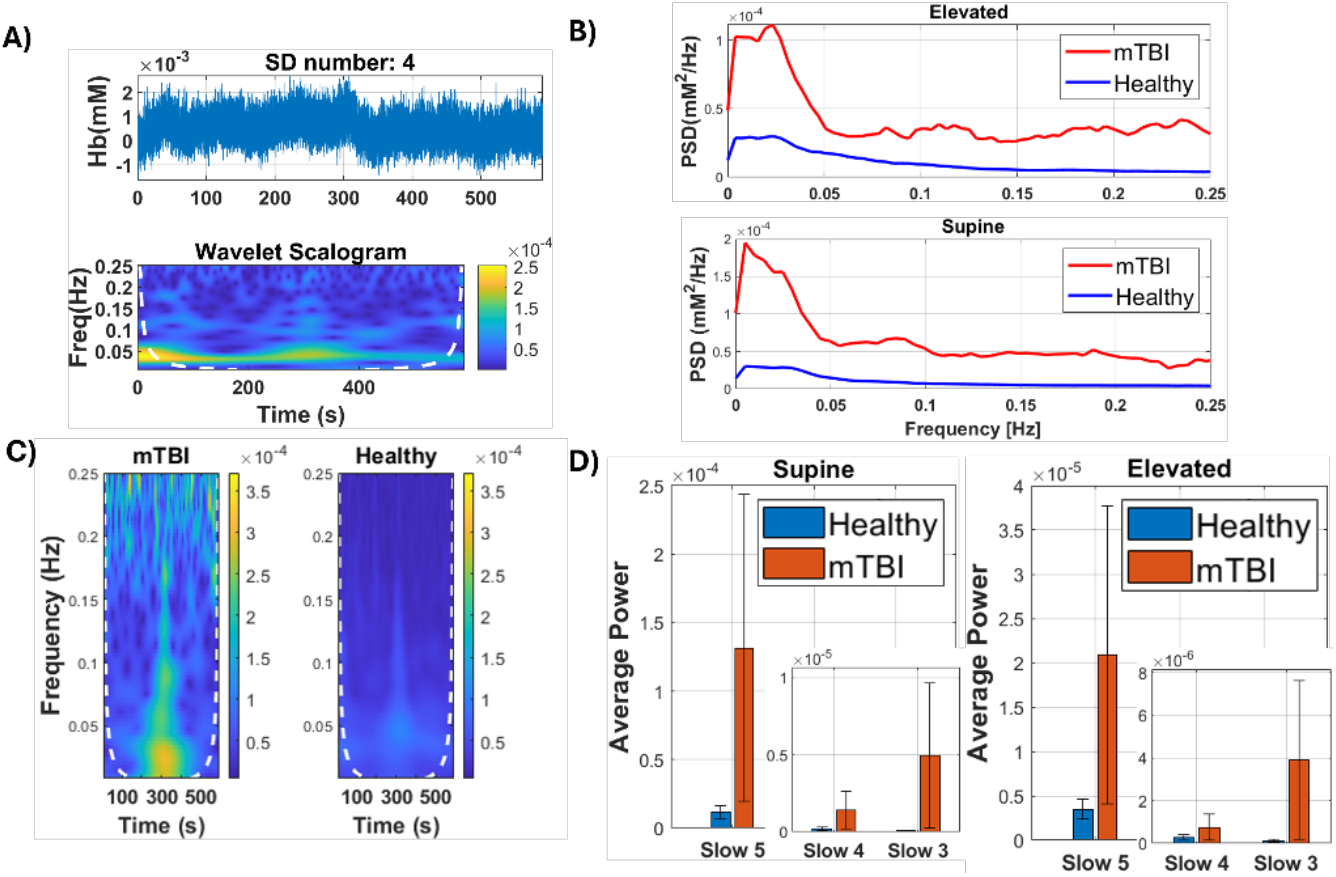
Analysis of Deoxy-Hemoglobin (Hb) concentrations in all subjects. (A) A single subject’s deoxyhemoglobin concentration (Hb) time series and the scalogram calculated from the continuous wavelet transform (CWT) are put side by side. This was done for every subject. (B) Healthy and mTBI elevated and supine positions are plotted in the frequency domain using fast Fourier transform. (C) The CWT plot shows a dashed white line showing the cone of influence (COI) that depicts areas where data interpretation is not accurate due to the stretching nature of wavelets. (D) Healthy and mTBI elevated and supine positions are plotted into summed bar plots processed using welch. Frequencies are grouped into slow bands, Slow 5: 0.01–0.027 Hz, Slow 4: 0.027–0.073 Hz, Slow 3: 0.073–0.198 Hz.

## 4. Discussion

This study aimed to identify hemodynamic differences between mTBI and healthy groups during positional changes. Through signal processing methods, FFT was used to maximize frequency resolution to identify any potential peaks of interest. However, spectral peaks seen in mTBI were most likely due to the small sample size of mTBI compared to healthy. Therefore, as an additional evaluation for a more reliable and robust PSD measurement. Welch’s method was applied to estimate the total power of LFOs during the steady-state conditions to show more accurate overall LFO PSD bar plots. Additionally, to observe any potential transient changes during the posture transition period, a continuous wavelet transform was used to observe frequency changes over time [54,55]. Our results suggest that mTBI subjects exhibit higher vasomotor responses due to postural stress, likely due to impaired cerebral autoregulation. mTBI had a larger change in HbO, Hb, and THC when going from elevated to supine. The higher changes in hemodynamic parameters could indicate mTBI’s cerebrovascular system’s lack in its ability to autoregulate. A similar outcome was seen in a study on HOB for TBI and healthy subjects where all changes in hemodynamic parameters were significantly higher in TBI compared to healthy [15]. Additionally, THC exhibited the largest differences between the two groups, this aligns with a previous study that found THC produced results that were more resilient to experimental noise and enabled the retention of more data [56]. However, some studies on TBIs reveal reduced HbO levels due to cognitive impairment. When exposed to targeted and contextual stimuli, TBI patients showed a weaker hemodynamic response than healthy controls [12]. Additionally, in stroke, atherosclerosis, and the aging populations LFO amplitude was lower, most likely due to vasomotor paralysis [39]. Our opposing results suggest a separate hemodynamic response when comparing mTBI environmental cerebral autoregulation effects to cognitive performance in brain injured cases and other neurological conditions. In the frequency domain, higher LFOs during the supine baseline were observed. This can be clearly seen in the CWT analysis where LFO power in mTBI is noticeably higher during and for a minute after the transition period compared to healthy. If the supine baseline itself were to be an indicator this high LFO power would be sustained throughout the CWT supine time. The distinctly high LFO power seen in mTBI in Slow-5 band throughout the entirety of the experiment could be an indicator of resting cerebral autoregulation impairments. During the transition period is where postural cerebral autoregulation impairments could be seen, as this is when the subjects are moved into a new position and the body would have to adjust for these positional changes. Other studies comparing diseased subjects to healthy have also seen this trend in higher LFOs [29,38,57–60]. This could potentially be a protective response, high LFOs reflect higher vasomotion to stabilize microcirculation. This response can help maintain oxygen delivery and perfusion during environmental changes, such as elevation adjustments. Based on these preliminary results, LFOs show promise as a biomarker for impaired cerebral autoregulation in mild TBI.

Limitations for this study were primarily due to its low statistical power from a limited sample size of mTBI. Another limitation is the absence of continuous mean arterial pressure (MAP) monitoring, which limits the ability to differentiate cerebral effects from systemic physiological reactions. Additionally, no short separation regression was used in this study due to ISS probe constraints, therefore the signals from the scalp and skull could not be mitigated [61]. Although the setup and protocol aimed to minimize movement, motion artifacts may still have contributed to elevated LFO power during positional transitions. However, the focus was on the difference between mTBI and healthy PSD magnitude, where no other healthy subject showed as large PSDs during that period. Additionally, a rigorous repeatable methodology was used to minimize any results from motion artifacts, as described in the methods. Further, this study relied on the assumption that hemodynamic changes were regional by placing the probe on the frontal lobe.

Machine learning approaches hold strong potential for improving diagnostic accuracy in mTBI by identifying frequency-specific biomarkers. Recent progress includes the application of deep learning models [63], for example, in reconstructing fNIRS signals [62], as well as the developing use of machine learning for fNIRS-based diagnostic applications [65]. Building on emerging potential of LFOs as a biomarker for mTBI, future efforts should explore automated classification models trained on expanded datasets to differentiate between healthy and mTBI subjects, or the severity of mTBI. These advancements will support the development of using diffuse optics modalities, like fNIRS and LFOs as an objective tool for mTBI diagnosis and real-time monitoring.

## 5. Conclusion

We propose that fNIRS-derived LFOs, especially THC-based measures in the Slow 5 (0.01–0.027 Hz) may serve as early indicators of cerebral autoregulation dysfunction in mTBI. Based on concentration, frequency, and time-frequency analysis mTBI subjects showed larger changes and magnitude in HbO, Hb, and THC. These differences may reflect impaired cerebral autoregulation in mTBI. THC demonstrated largest magnitude differences, supporting prior findings of THC’s robustness. Elevated LFOs in mTBI may indicate a dysfunctional or overcompensated vasomotor response to positional stress. These findings align with prior work linking irregular LFOs to injuries or conditions that would lead to dysregulated autoregulation. This work emphasizes the potential of LFO analysis as a noninvasive, frequency-based marker for detecting mTBI.

## Funding

NIH R01 (NIBIB Brain Initiative, 7R01EB031759-03).

## Acknowledgment

We would like to thank Sahar Sabaghian and Irfaan Dar for their signal processing guidance and expertise.

## Disclosure

The authors declare no conflicts of interest.

## Data availability

Data underlying the results presented in this paper are not publicly available at this time but may be obtained from the authors upon reasonable request.

## Notes

### Competing Interest Statement

The authors have declared no competing interest.

### Funding Statement

The study was funded by NIH R01 (NIBIB Brain Initiative, 7R01EB031759-03).

### Author Declarations

The Instutional Review Board of Wright State University approved this protocol.

